# Status Quo and Influencing Factors of Family Doctors Performance in Dongguan City

**DOI:** 10.1101/2021.09.23.21263974

**Authors:** Jiao Wang, Junhui Xiao, Xiali Li, Lin Zhang, Na Wang

## Abstract

**Background:** The low-performance rate of family doctors is the most important problem in the implementation of the contract service system of family doctors in China, and improving the performance rate of family doctors has become a topic of common concern. We investigated the performance behavior of family doctors in Dongguan, Guangdong Province, to understand the current situation of family doctors performance in Dongguan, and to explore the influencing factors of their performance.

**Objectives:** To understand the status quo of family doctors performance in Dongguan City, Guangdong Province, and to explore the influencing factors of family doctors agreement fulfillment rate.

**Methods:** The multi-stage sampling method was used to randomly select family doctors in Dongguan City, Guangdong Province, to investigate the status quo of family doctors performance in community health service institutions in Dongguan City. Logistic regression analysis was used to investigate the influencing factors of family doctors performance, and decision trees and associations were used to further identify the influencing factors of family doctors performance.

**Results:** 100 family doctors participated in the survey, and 75% of them were generally able to perform their duties. Logistic regression analysis shows that whether a family doctor signs a contract with a resident is the influencing factor of family doctor performance behavior, and the higher the family doctor signing rate, the higher the family doctor performance rate. The results of decision tree analysis showed that the biggest influence factor of family doctors performance behavior is whether the family doctors are contracted residents or not. ROC curve was 0.860, and pAUC was 0.073 respectively. The most influential factor in decreasing the average Gini index of random forest is whether the family doctor signed a contract or not. The results of association rules showed that there was a correlation between whether the family doctor signed a contract or not and the performance of the family doctor.

**Conclusions:** The performance rate of family doctors in Dongguan City, Guangdong Province is relatively high. Whether family doctors sign contracts with residents is the influencing factor of family doctors performance. Improving the contract rate of family doctors is the key to improve the performance rate of family doctors.

## Introduction

Family doctor contract system refers to a service system in which a general practitioner establishes a long-term therapeutic relationship with community members through contract signing to manage the health need of the clients. Implementing the family doctor contract system can facilitate the residents’ access to medical services and healthcare needs. It is an essential means to alleviate the health economic burden of the residents^[1,2]^

The family doctor contract system is a widely implemented diagnosis and treatment system in more than 50 countries or regions around the world^[3]^. China established the family doctor contract system in 2009. Many opinions and suggestions on the family doctor contract system have been voiced since, and the strategic position of the family doctor contract system in the new medical reform has been gradually revealed. Under the current situation, China’s new medical reform has entered the deep water zone, and the family doctor contract system has become an important direction to consolidate the medical reform.

By the end of November 2017, the number of the resident who signed the contract with family doctors in China has exceeded 500 million, and the signing coverage rate of the population has exceeded 35%^[4]^. At present, the signing rate in pilot areas in China has even reached 47.6%^[5]^. Although the implementation of the contract system for family doctors in China has achieved certain success, some family doctors signed the contracts with residents without providing substantial health service, which does not benefit the contracted residents. Therefore, the family doctor contract system was not implemented effectively. From this situation, the performance rate of family doctors become an important index to measure the quality of care from family doctors contract.

According to the data of the Health Statistics Yearbook of China, the number of general practitioners in 2019 was 365,082 (the data came from the 2020 China Health Statistics Yearbook), and only some of these family doctors were able to perform the contracted services. Based on the data of 500 million contracted residents in China, the number and frequency of contracted services for family doctors per capita can be very limited. Therefore, the issues of performance behavior of family doctors, such as low-performance rate or “signing without agreement”, poor performance service ability of family doctors, and single performance service, are exposed. Among the above issues, the most urgent problem to be solved is the low compliance rate of family doctors. At present, the compliance rate in China is only 37.2%^[6]^, which indicated that the overall situation is not optimistic. If the problem of low performance rate of family doctors is not solved as soon as possible, the contract system of family doctors cannot develop sustainably.

Dongguan City, Guangdong Province, is one of the cities where the family doctor contract system was implemented earlier in China^[7]^. By 2020, Dongguan City had 114 teams of family doctors who are experienced in the practice of contract service, which is representative and can be used as a reference to study the contract service and performance of family doctors in China. There are relevant literatures on family doctor’s contract service in China. However, there are few studies on family doctors’ performance behavior. Although some scholars have clarified the current situation of family doctors’ performance behavior, they have not explored the causes from the perspective of family doctors. This study intends to clarify the behavior and motivation of family doctors’ performance through the field investigation of family doctors’ performance behavior in Dongguan City, Guangdong Province. Therefore, the objectives of this study are to understand the status quo of family doctors’ performance and to explore the influencing factors of performance behavior of family doctors in Dongguan City, Guangdong Province.

## Method

### Population

The questionnaire survey of family doctors’ performance behavior was supported and affirmed by Dongguan Government and Health Service Institutions in Guangdong Province, and all the respondents obtained informed consent. From March 2020 to August 2020. 100 family doctors who work in 24 community health service institutions in eight towns (or districts) of Dongguan participated in the study. There were 14 people in Dongcheng, 15 in Shilong, 12 in Chang’an, 15 in Songshan Lake Industrial Park, 13 in Dalang, 11 in Dalingshan, 10 in Dongkeng and 10 in Liaobu.

### Study variables

The questionnaire variables of this study mainly included basic demographic data (sex, age, education background, professional certificate, monthly income, years of work at the grass-roots level), the awareness of the Guangdong Family Doctor Service Package, the attitude towards the government’s implementation of family doctor service, whether one’s ability can meet the requirements of family doctor service, and whether to sign a family doctor with residents, the type of service, number of family doctors signing contracts, number of family doctors participating in training within one year, performance behavior of family doctors and other related information.

### Research method

The multi-stage sampling method was used to select the research participants. Firstly, according to the Administrative Division of Dongguan City, Guangdong Province, eight towns (or districts) were selected by simple random sampling method, and three community health service institutions were randomly selected in each town (or district) as well. Therefore, the number of family doctors’ team members was based on the numbering basis in each community health service institution. Five family doctors (or team members) were randomly selected by equidistant random sampling method, and the performance behavior of family doctors was investigated. A total of 120 family doctors (or team members) were from 24 community health service institutions.

### Data collection and collation

After obtaining the informed consent of each participant in advance, the participants were investigated based on the survey. After the investigators introduced the relevant information, the participants filled out the questionnaire anonymously and the survey was collected on the spot. A total of 120 questionnaires were distributed with a recovery rate of 100%. 100 valid questionnaires were obtained, and the effective rate was 83.33%. Unified numbering of valid questionnaires was applied.

### Data analysis/statistical analysis

The data were analyzed by SPSS22.0 statistical software and R package, and the frequency distribution of each index was understood by statistical description. Chi-square test and logistic regression analysis were used to analyze the influencing factors of family doctors’ performance behavior in community health service institutions, and decision trees and association rules were used to further find out the influencing factors of family doctors’ performance behavior.

## Result

### Basic information of respondents

The demographic information of 100 family doctors in community health service institutions was obtained and shown in **Table 1**. Among them, most family doctors are 20-45 years old, with college education, junior and intermediate titles, more than 5 years working experience, and 5,000-9,000 yuan monthly income.

**Table 1.** Demographic information of Family Doctors in this study.

| Variable |  | Number of Participants | Frequency (%) |
| --- | --- | --- | --- |
| Sex | Male | 42 | 42.00 |
|  | Female | 58 | 58.00 |
| Age | 20-35 years old | 43 | 43.00 |
|  | 35-45 years old | 32 | 32.00 |
|  | ≥45 years old | 25 | 25.00 |
| Academic degree | Secondary school and below | 13 | 13.00 |
|  | Universities and Colleges | 73 | 73.00 |
|  | Master degree or above | 14 | 14.00 |
| Professional certificate | Primary | 33 | 33.00 |
|  | Secondary | 46 | 46.00 |
|  | Senior | 21 | 21.00 |
| Years of grass-roots work | ≤2 years | 5 | 5.00 |
|  | 3-5 years | 16 | 16.00 |
|  | 5-10 years | 31 | 31.00 |
|  | ≥10 years | 48 | 48.00 |
| Monthly income | ≤3000 yuan | 4 | 4.00 |
|  | 3000-5000 yuan | 16 | 16.00 |
|  | 5000-7000 yuan | 37 | 37.00 |
|  | 7000-9000 yuan | 24 | 24.00 |
|  | ≥9000 yuan | 19 | 19.00 |

### Analysis of family doctors’ signing and performance behavior

A statistical analysis of the related indicators of family doctors’ contract signing and performance showed that most family doctors have already known the contents of Guangdong Family Doctor Service Package, and 60% considered themselves as competent for family doctor service at the current professional level. 82% of family doctors have signed family doctor service with residents.62% of the family doctors signed contracts with 90 households or above, and most of them participated in the training of family doctors less than six times a year, and 75% of the family doctors were able to perform the contract service for family doctors, as seen **Table 2**.

**Table 2.** Contract signing and performance behavior of family doctors.

| Variable |  | Number of Participants | Frequency (%) |
| --- | --- | --- | --- |
| Do you know the Family Doctor Service Package of Guangdong Province | yes | 99 | 99.00 |
|  | no | 1 | 1.00 |
| Can you meet the requirements of family doctor service | yes | 60 | 60.00 |
|  | no | 40 | 40.00 |
| Have you signed a family doctor service with residents | yes | 82 | 82.00 |
|  | no | 18 | 18.00 |
| Number of contracts signed by family doctors | Under 90 households | 38 | 38.00 |
|  | 90-120 households | 21 | 21.00 |
|  | More than 120 households | 41 | 41.00 |
| Number of trainings attended by family doctors in one year | Twice and below | 43 | 43.00 |
|  | 3-5 times | 44 | 44.00 |
|  | 6 times and above | 13 | 13.00 |
| Do family doctors perform their duties | Completely able to | 12 | 12.00 |
|  | Able to | 30 | 30.00 |
|  | common | 33 | 33.00 |
|  | Seldom able to | 7 | 7.00 |
|  | Unable to | 18 | 18.00 |

### Logistic regression analysis of family doctors’ performance behavior

#### Univariate analysis

In the analysis of performance indicators of family doctors, the univariate analysis was carried out with the classification criteria of complete ability, basic ability, general ability, a small number of options, and complete inability. The expected value is less than 5 or more than 20% of the cells with the expected value less than 5 appear in the analysis results, and the low-frequency options affected the fitting of the Logistic regression model. Therefore, by adjusting the classification standard of family doctors’ performance behavior, it was completely and able to be merged into family doctors’ performance behavior (i.e. =1), and a small number of them were merged into family doctors’ non-performance behavior (i.e., no =0).

With the performance behavior of family doctors (yes =1, no =0) as the dependent variable, sex, age, educational background, professional certificate, monthly income, working years, whether know the Family Doctor Service Package of Guangdong Province, attitude towards the government’s implementation of family doctor service, whether agree with the family doctor’s first consultation responsibility system, whether agree with the general practitioner teamwork system, whether can meet the requirements of family doctor service, whether sign a family doctor with residents. 14 factors, such as type service, the number of family doctors signing contracts, and the number of family doctors participating in family doctor training in one year, were identified as independent variables, and the influencing factors of family doctors’ performance behavior were analyzed by univariate analysis. According to the results of univariable analysis, whether or not to sign family doctor service with residents, the number of contracts signed by family doctors, and the number of family doctors participating in family doctor training in one year have an influence on the performance behavior of family doctors (P<0.05), as seen in **Table 3**.

**Table 3.**
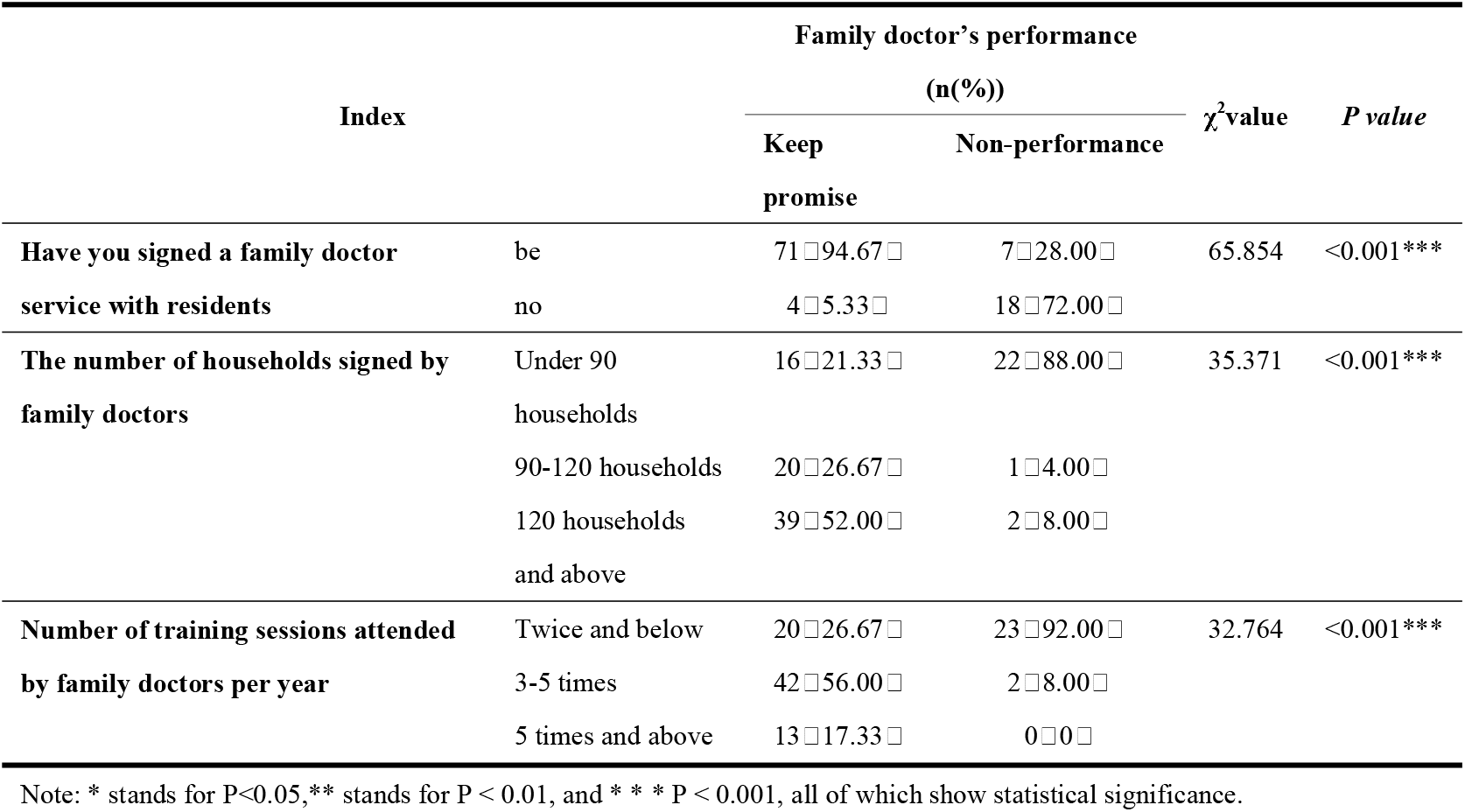
Univariate analysis results of influencing factors of family doctors’ performance behavior.

### Multivariate analysis

After screening by univariate analysis, the performance behavior of family doctors (yes =1, no =0) was identified as a dependent variable, and the three possible influencing factors (whether to sign family doctor service with residents, the number of contracts signed by family doctors, and the number of family doctors participating in family doctor training in one year) were defined as independent variables for binary logistic regression analysis, as seen in **Table 4** for a description of assignment of variables. Chi-square test P value and pseudo R^2^ in the model fitting information are P<0.001 and P=0.542, indicating that the final model fitting was better.

**Table 4.** Explanation of variable assignment of influencing factors of performance behavior of family doctors.

| Variable name | Define assignment |
| --- | --- |
| Have you signed a family doctor service with residents | Yes =1, no =2; |
| Number of contracts signed by family doctors | Under 90 households =1,90-120 households =2,120 households and below =3; |
| Number of training attended by family doctors in one year | Twice and below =1,3-5 times =2, 6 times and above =3. |

The results of binary logistic regression analysis showed that whether to sign family doctor service with residents was an influencing factor of family doctor’s performance behavior, and the higher the family doctor’s signing rate, the higher the family doctor’s performance rate, as seen in **Table 5**.

**Table 5.** Bivariate Logistic Regression Analysis of Family Doctors’ Performance Behavior.

| Index | *B* | *SE* | *Wals* | *P* | *OR* |
| --- | --- | --- | --- | --- | --- |
| Have you signed a family doctor service with residents | -22.093 | 0.505 | 3.437 | 0.045* | 2.434 |

Note: \* means $P<0.05$ , indicating that the difference is statistically significant.

### Decision tree analysis of family doctors’ performance behavior

Decision tree is a supervised learning method, which can summarize decision rules from a series of data with features and labels to solve the problems of classification and regression^[8]^. In this research, family doctors’ performance behavior was also analyzed by decision tree, and CHAID’s decision tree growth algorithm was adopted. In addition, this study used the split sample verification method to verify. 70% (70 samples) were randomly selected as training samples and the remaining 30% (30 samples) as test samples from the data set (100 samples). The dependent variable was family doctor’s performance behavior, and the independent variable was whether to sign family doctor’s service with residents, the number of family doctors signing contracts, and the number of family doctors participating in family doctor training in one year. The output model summary showed in **Table 6**.

**Table 6.** CHAID Model Summary.

| Specify item | Growth method | CHAID |
| --- | --- | --- |
|  | dependent variable | Performance behavior of family doctors |
|  | independent variable | Sex, age, educational background, professional title, monthly income, working years at the grass-roots level,<br>Do you know the Family Doctor Service Package of Guangdong Province?<br>Attitude towards the government's implementation of family doctor service,<br>Do you agree with the family doctor's first consultation responsibility system?<br>Do you agree with the teamwork system of general practitioners?<br>Can you meet the requirements of family doctor service,<br>Whether to sign family doctor service with residents,<br>Number of family doctors signing up,<br>Number of family doctors participating in family doctor training in one year |
|  | test and verify | Split sample |
|  | maximum depth | 3 |
|  | Minimum case in the parent node | 5 |
|  | Minimum case in the child node | 2 |
| **Result** | Included arguments | Do you sign a family doctor service with residents? |
|  | Number of nodes | 3 |
|  | The final number of nodes | 2 |
|  | depth | 2 |

The results of decision tree analysis show that the tree structure is only divided into one layer, that is, it is divided according to whether the family doctor signed a contract with residents. Therefore, the dominating factor affecting the family doctor’s performance behavior is whether the family doctor signed a contract with residents, which shows that whether the family doctor signed a contract became an important indicator for the family doctor’s performance behavior as seen in **Figure 1**.

**Figure 1.**
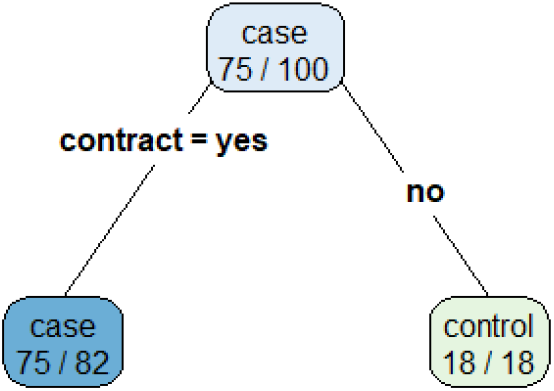
Decision tree model of family doctor’s performance behavior.

As an example to better illustrate the decision tree model, this paper applies the confusion matrix and ROC curve to evaluate the corresponding model. See **Table 7** for the confusion matrix.

**Table 7.** Confusion matrix of decision tree model of family doctor’s performance behavior.

| Prediction | Absent present |  |
| --- | --- | --- |
|  | no | yes |
| no | 8 | 0 |
| yes | 3 | 19 |

Based on the confusion matrix, the classification accuracy, sensitivity and specificity of this data can be obtained as follows:

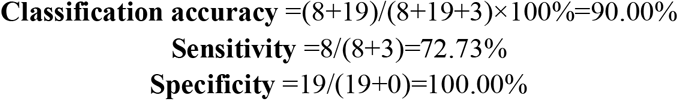

The classification accuracy of the family doctor’s performance model was 90%, and the sensitivity and specificity were both high. Therefore, the decision tree model of the family doctor’s performance behavior was better.

ROC curve is the “receiver operating characteristic” curve. ROC curve analysis can test the actual positive and negative abilities^[9]^. AUC (Area Under Curve) is the area under the ROC curve, which is between 0.1 and 1. It can be used to evaluate the classifier. The larger the AUC value was, the fitting of the classifier was better^[10]^.

The data analysis results of the decision tree model of family doctors’ performance showed that the area value under the ROC curve of the decision tree model was 0.860, and the 95% confidence interval was from 0.799 to 0.909, which indicated that the diagnosis of family doctors’ performance behavior was effective. This also supported the results of Logistic regression analysis as seen in **Figure 2**.

**Fig. 2.**
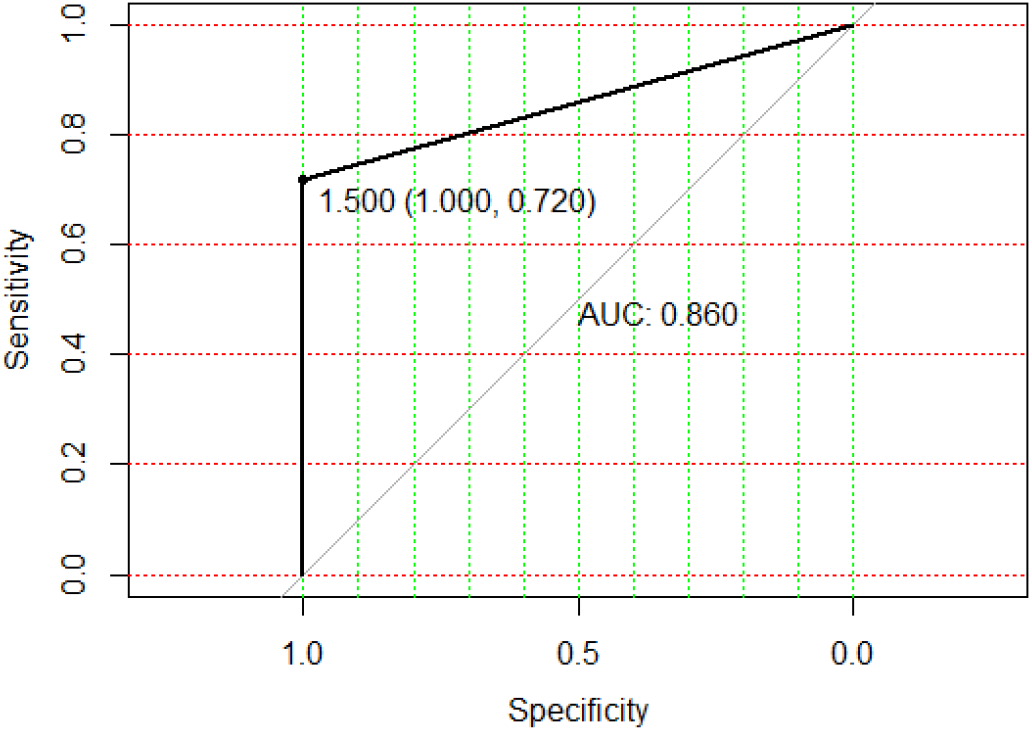
ROC curve of decision tree model of family doctor’s performance behavior.

When there were relatively few positives and negatives in the data, we need high specificity to avoid a large number of false positives, and ideally, we need high-sensitivity cooperation. AUC is the area under the whole specificity or sensitivity level, while pAUC is the AUC obtained by paying attention to a certain sensitivity or a certain specificity. Especially, the ROC curve with high sensitivity or specificity has a good evaluation effect^[11]^. In this study, the specificity is 100.00%, so we need to pay attention to AUC with the specificity of 90%-100%, and get the ROC curve (pAUC) with a specificity ≥90%. Therefore, we need to analyze the pAUC value of the classifier evaluation index with the specificity of 100%. The AUC in the range of 90%-100% specificity provided a vertical perspective, the total AUC is 0.1, while the pAUC is 0.073 which indicated that whether the family doctor signed the contract or not has a good explanation for the performance behavior of the family doctor, which was consistent with the results of ROC curve analysis, as seen in **Figure 4** for analysis of pROC curve.

**Fig. 3.**
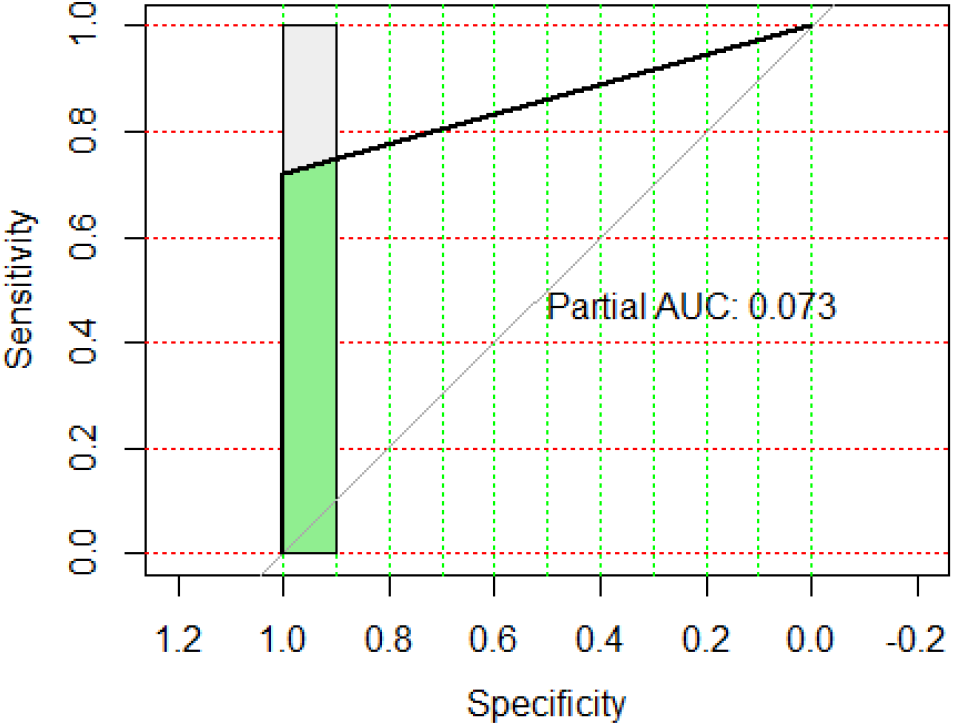
ROC curve of decision tree model of family doctor’s performance behavior in a specific state.

**Fig. 4.**
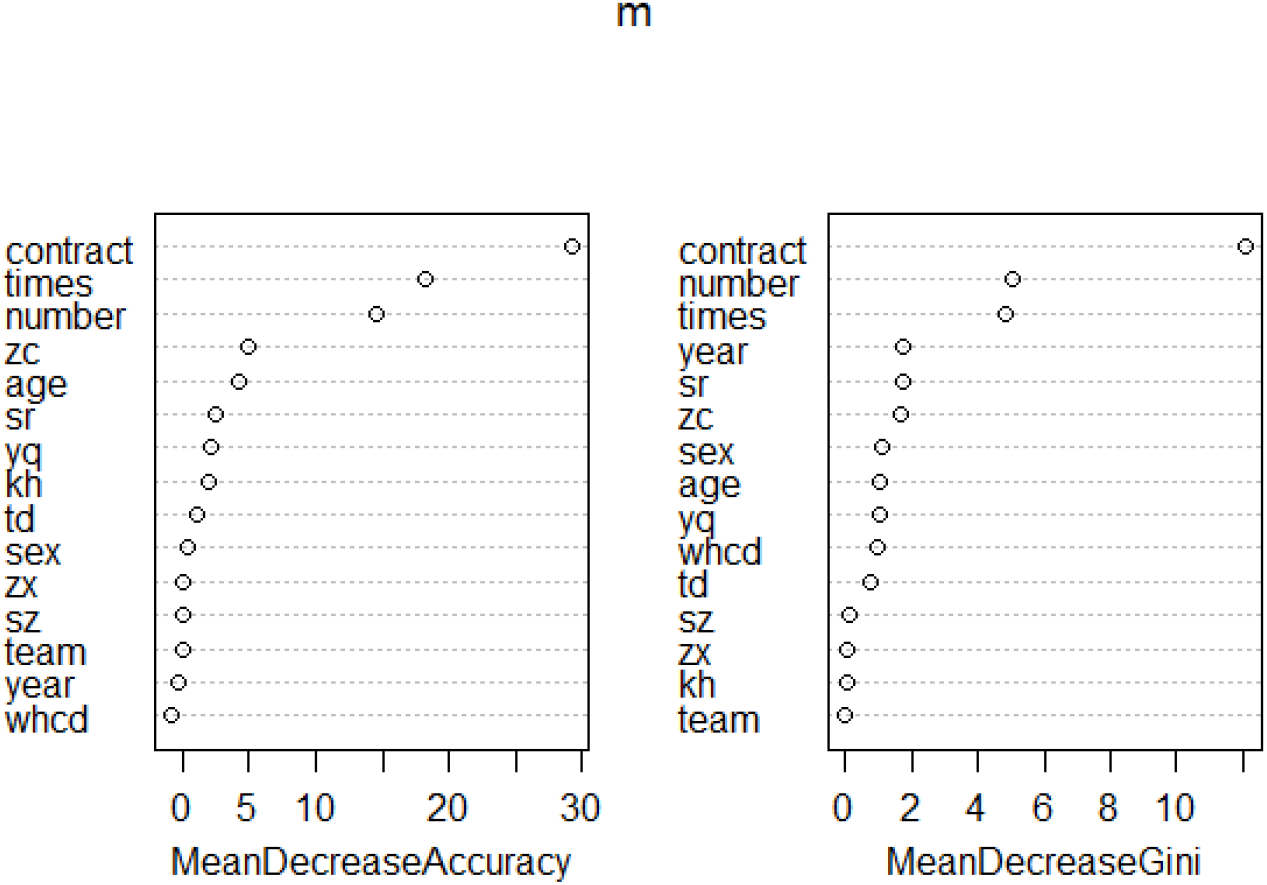
Feature attributes of random forest extraction.

From the results of improving the accuracy of random forest and node impurity, it can be seen that the most important characteristic variable that affects the performance behavior of family doctors is whether or not they sign a contract. In the feature attribute map of random forest extraction, the greater the decrease of average accuracy and average Gini index, the more impactful the variable. This variable had an impact on the performance behavior of family doctors. From the results of feature attributes extracted from random forest, it can be seen that whether family doctors sign up as residents was an important influencing variable of family doctors’ performance behavior, as shown in **Table 8** and **Figure 4**. Based on the confusion matrix, we can see that the classification error rate (OOB)=3/(8+19+3)=10%, which shows that the random forest model is better. This is consistent with the results of the decision tree model.

**Table 8.** Average accuracy reduction and average Gini index reduction of each index.

| Variable | Average accuracy reduction | Average Gini index decrease |
| --- | --- | --- |

|  | (Mean Decrease Accuracy) | (Mean Decrease Gini) |
| --- | --- | --- |
| contract | 29.3498 | 12.1039 |
| times | 18.2522 | 4.8497 |
| number | 14.4795 | 5.0719 |
| age | 4.1863 | 1.0740 |
| sex | 0.4385 | 1.1398 |
| whcd | -0.8318 | 0.9627 |
| zc | 4.9906 | 1.6401 |
| year | -0.2517 | 1.7464 |
| Sr | 2.5301 | 1.7408 |
| zx | 0.0000 | 0.0838 |
| td | 1.1283 | 0.7649 |
| sz | 0.0000 | 0.1172 |
| team | 0.0000 | 0.000 |
| kh | 1.8752 | 0.0832 |
| yq | 2.1666 | 1.0150 |

## Discussion

### Signing rate

Since the implementation of the family doctor contract system in China in 2009, many national policies have gradually revealed the importance of the family doctor contract system in the healthcare system, and further consolidate the development direction of the family doctor contract system in China. Under the guidance of relevant policies, China’s family doctor signing system has achieved gratifying results so far. As of the end of November 2017, The number of contracted doctors in China has exceeded 500 million, and the contracted coverage rate of the population has exceeded 35.0%^[4]^, which is the landmark cut-off data for the success of the initial trial of Chinese family doctor contracting service. By 2019, the contracted rate in China’s pilot areas has reached 47.6%^[5]^. The signing rate of family doctors in Zhejiang Province surveyed by Qiu□ YW^[12]^ is 50.43%. From our research results, we can see that the signing rate of family doctors in Dongguan City, Guangdong Province is considerable, which has exceeded the national and provincial level of the signing rate of family doctors.

Guangdong Province is one of the provinces where the family doctor contract system was implemented earlier in China^[13]^. Since 2009, provincial and municipal government departments have issued several policies related to the family doctor contract system. In 2018, Dongguan formulated the Implementation Plan for Promoting the Family Doctor Contract Service System in Dongguan (Draft for Soliciting Opinions) ^[14^. Breakthroughs have been made in the ways, contents, collection and payment, performance appraisal, and incentive mechanism of family doctors’ contract signing, which promotes the construction of family doctors’ contract signing service. In China, the implementation of government policies plays a positive and important role in the signing of family doctors^[15]^ □ Dongguan is not unique in its effort to implement the family doctor signing system, which is a powerful guarantee for the high signing rate of family doctors in Dongguan. In addition, the government and relevant departments have boosted the publicity^[16,17]^. Multi-channel and multi-dimensional publicity of the family doctor signing system^[16,18]^ is an effective driving force for the high signing rate of family doctors in Dongguan. Finally, under the call of the policy, community health service institutions set up a family doctor team, which was attended by family doctors, nurses, pharmacists, and Chinese medicine practitioners. This is the core element of Dongguan family doctor contract system. The family doctor team focuses on active service, classifies the population according to the health status and health needs of residents, signs different service packages, and provides targeted family doctor signing services for different categories of subgroups. It mainly includes basic medical services, basic public health services, and health management. Based on the above reasons, the family doctor signing service in Dongguan can maintain a high signing rate. By the end of 2020, 2.68 million people in Dongguan had signed family doctors, with a total of 2.15 million family doctor service packages and 160,000 fee packages^[19]^.

### Agreement fulfillment rate

The starting point of family doctor’s contract service is the high rate of family doctor’s contract, and the essence of family doctor’s contract service is its high-performance rate. Only when the contracted patients achieve a high-performance rate can they promote the sustainable process of the family doctor contract service and realize the original purpose of the family doctor contract service. At present, the performance of family doctors in China is suboptimal^[20]^. The most prominent phenomenon is the problem of family doctors signing without agreement^[4,21]^, that is, the performance rate of family doctors is low. The research data of Jie Chen^[6]^ shows that the performance rate of family doctors in China in 2016 was only 37.2%. In this survey, the compliance rate of family doctors in Dongguan City, Guangdong Province is as high as 75.0%, which is higher than that of Jie Chen^[]^.

Such a high-performance rate of family doctors is closely related to the family doctor team implemented in Dongguan. At the beginning of the implementation of the family doctor contract system in China, Dongguan began to set up a family doctor team, regularly training the family doctor team members every year, informing them of policy trends and performance duties, etc., so that every family doctor can know this task. Each community health service institution will set up a team of 2-3 family doctors, and mobilize the family doctor contracting service of community health service institutions through cooperation and coordination among the family doctor teams^[16]^.

This is an effective way to improve the performance rate of family doctors. Each family doctor team will establish effective communication channels with the contracted residents, such as WeChat and telephone. For example, in the WeChat group of a family doctor team, the family doctor will publish some health knowledge either regularly or sporadically, and the family doctor will provide corresponding medical services for the problems raised by the contracted residents to ensure timely and effective medical services^[22]^. The family doctor will also prompt the contracted residents to utilize the family doctor contract service through on-site appointments or telephone appointments, which is very helpful for patients with chronic diseases such as hypertension and diabetes. Of course, poster propaganda also played a certain auxiliary role.

### The signing rate is an important factor affecting the performance rate

Logistic regression analysis of this study shows that only residents who sign contracts are the influencing factors of family doctors’ performance, and the higher the signing rate of family doctors, the higher the performance rate. Decision tree model and random forest model are classified statistical analysis methods, which can extract important feature attributes from massive indexes. Therefore, decision tree model and stochastic forest model can analyze the present situation from another perspective and can be regarded as a supplement to Logistic regression model when analyzing statistical results. The results of decision tree model and stochastic forest model show that whether a family doctor signs a contract with a resident is a factor that affects the performance behavior of family doctors. When the signing rate of family doctors is higher, the higher compliance rate of family doctors. The analysis results of various models are highly consistent, which once again supports the conclusion that whether family doctors sign up for residents’ service behavior is the biggest influencing factor of family doctors’ performance behavior. Therefore, we can understand that if we want to improve the performance rate of family doctors, we should first improve the contract rate of family doctors.

And what is the signing rate of family doctors in China? The research in Jie Chen^[6]^ shows that the signing rate of family doctors in China’s pilot areas has reached 47.6%, compared with the United Kingdom^[23]^, the United States^[24]^, and Germany^[25]^, the contract performance of family doctors in China is still low. This is related to the development of China’s family doctor signing system. Huang JL^[26]^ believes that the four basic elements of the establishment of the Chinese family doctor system are still relatively weak, and the family doctor signing rate will be affected to a certain extent. The family doctor contract system in China was established in 2009. In 2011, the State Council issued Guiding Opinions on Establishing General Practitioner System, in 2016, the Guiding Opinions on Promoting Family Doctor Contract Service, and in 2019, From the perspective of policy constraints, the Notice on Doing a Good Job of Family Doctor Contract Service in 2019 gradually clarified the important position of the family doctor contract system in China’s new medical reform. And further deepened the development direction of the family doctor contract system. Under the current situation, the family doctor contract system has become an important direction to deepen the new medical reform. Under such policy guidance, the signing rate of family doctors can reach nearly 50.0%, which is already a gratifying achievement.

It is precise because of the low contracting rate of family doctors in China that the phenomenon of “signing but not contracting” occurs. This is the most prominent problem in the signing and performance services of family doctors in China at this stage ^[4]^. The lower th signing rate, the lower the compliance rate of family doctors, which can explain the above phenomenon. Increasing the rate of family doctors signing is the most effective solution to tackle the outstanding problem of some family doctors “signing but not contracting”. Combining the current status of family doctor performance services by many scholars ^[12,27,28]^ and the results of this study, we can make a reasonable inference: the low-performance rate of family doctors and “signing but not contracting” problems are the initial stage of family doctors signing services When the phenomenon of widespread contracting by family doctors is formed, problems such as the rate of performance of family doctors and “signing but not contracting” will be solved.

The results of this study show that the high contract rate of family doctors leads to high-performance rate. It can be seen that Dongguan City, where the contract rate of family doctors is as high as 82.0%, has laid a good foundation for the performance behavior of family doctors in this area, and the high contract rate of family doctors is the primary guarantee for the high-performance rate. In addition, high performance rate of family doctors in Dongguan can not be separated from the support of the policy environment of Dongguan family doctors’ contract signing system and the establishment of family doctors’ awareness of performing duties. Since the establishment of the family doctor contract system, Dongguan has publicized the family doctor contract system utilizing multi-channel and multi-ways, in which the effect of on-site contract signing is obvious^[16^]. On-site signing refers to signing a family doctor and service team directly with a patient when he visits a community health service institution, which is fast and efficient. During the on-site signing process, the contracted residents will enter the WeChat group of the family doctor team through the WeChat platform, and this group will send medical service information regularly. Thereby improving the performance rate of family doctors. At the same time, the establishment of family doctors’ team has enhanced their awareness of performing their duties. There are 114 family doctors’ teams in Dongguan so far, among which it is not uncommon to be awarded the title of National Excellent Family Doctors’ Team, providing residents with family doctors’ signing services of classified signing, paid signing, and differentiated signing. The family doctor team established and growing up in such an environment has a strong sense of performance responsibility, and will naturally have higher performance behavior.

### Conclusion

Our research findings are helpful for the government to formulate corresponding policies. The government may realize that the signing rate of family doctors is the major factor that affects the performance behavior of family doctors.

## Data Availability

Data Availability Statement The data that support the findings of this study are available on request from the corresponding author. The data are not publicly available due to privacy or ethical restrictions.

## Acknowledgments

This study was supported by the School of Humanities and Management, Guangdong Medical University.

